# A Novel Correction Method for QT Interval in the Presence of Left Bundle Branch Block Morphology

**DOI:** 10.64898/2026.06.16.26355834

**Authors:** Mohammad Alasti, Amin Esmailian, Colin Machado, Jagat Adhikari, Sing Huey Cheng, Francis Ha, Jeffrey Alison, Andrew Krahn, Hui Chen Han

## Abstract

**Background:** Accurate assessment of the QT interval is challenging in the presence of QRS prolongation, such as during ventricular pacing or bundle branch block. Current correction methods are heterogeneous and lack consensus. To evaluate the relationship between QRS duration and QT interval during ventricular pacing and to develop a practical correction method for QT assessment.

**Methods:** In this prospective single-centre study, 94 patients undergoing electrophysiology study for supraventricular tachycardia were included. Standardised pacing was performed at the same cycle length from the right ventricular (RV) apex, high output and low output pacing from His catheter, and coronary sinus (reference). QRS and QT intervals were measured from 12-lead ECGs. Changes in QT (ΔQT) and QRS duration (ΔQRS) were analysed using linear regression and mixed-effects modelling. QT correction formulas of the form QT corrected = QT − N × ΔQRS were evaluated using Bland–Altman analysis across multiple coefficients.

**Results:** A significant positive correlation between ΔQRS and ΔQT was observed across all pacing sites (r = 0.52–0.74, p < 0.001). In mixed-effects modelling, ΔQRS was a strong independent predictor of ΔQT (β ≈ 0.59, p < 0.001), with no significant interaction between pacing site and ΔQRS, supporting a consistent relationship across pacing locations. Bland–Altman analysis demonstrated that correction coefficients of 0.65–0.70 minimised systematic bias compared with lower coefficients, with similar precision across models (SD ≈ 16 ms) and no evidence of proportional bias. A coefficient of 0.65 provided the most balanced performance between bias and variability.

**Conclusion:** QT prolongation during ventricular pacing is primarily driven by QRS widening and follows a consistent linear relationship across pacing sites. A simple correction using QT corrected = QT − 0.65 × (QRS − 100 ms) provides a practical and accurate method for QT assessment, with potential clinical applicability in patients with conduction abnormalities or ventricular pacing.

## Introduction

Evaluation of the QT interval is a fundamental component of clinical practice for identifying patients at increased risk of ventricular arrhythmias and sudden cardiac death. This is particularly important in conditions such as long QT syndrome and in the presence of contributing factors including QT-prolonging medications, bradycardia, and electrolyte disturbances such as hypokalaemia. Several commonly prescribed antidepressants and antipsychotic agents have been associated with QT prolongation; therefore, electrocardiographic (ECG) monitoring and QT interval assessment are recommended when initiating these therapies in patients with underlying cardiac risk factors.^1,2^

Accurate assessment of the QT interval becomes challenging in the presence of QRS prolongation, such as bundle branch block (BBB) or ventricular pacing. Both right bundle branch block (RBBB) and left bundle branch block (LBBB) increase QRS duration and consequently prolong the measured QT interval, despite part of this prolongation reflecting delayed depolarization rather than true repolarization abnormalities.^3^ As a result, the QT interval requires adjustment to account for the contribution of QRS widening. Multiple correction methods have been proposed; however, no consensus has been reached regarding the optimal approach.^3–5^ Consequently, there remains no standardized method for QT interval assessment in patients with BBB or during right ventricular (RV) pacing.^6,7^

Among conduction abnormalities, LBBB has received particular attention because it is frequently encountered in patients with structural heart disease and is independently associated with increased all-cause mortality. In addition, RV pacing produces a widened QRS complex with an LBBB-like morphology, resulting in prolongation of the measured QT interval through extension of the depolarization phase. Ventricular pacing also alters the spatial and temporal sequence of ventricular activation, potentially influencing repolarization dynamics beyond QRS widening alone.^3^

Accordingly, numerous approaches have been proposed to correct the QT interval in the setting of LBBB and ventricular pacing. These methods generally involve subtracting either a fixed value or a proportion of the QRS duration from the measured QT interval, or modifying the QT measurement itself to account for delayed depolarization.^3–17^ Despite these efforts, currently available correction formulas demonstrate variable performance and limited agreement across different pacing and conduction patterns.

Therefore, the aim of this study was to evaluate the relationship between QRS duration and QT interval during ventricular pacing from different pacing sites and to develop a practical correction method for QT assessment in patients with LBBB or ventricular pacing.

## Materials and Methods

### Study Design

This was a single-centre, prospective cohort study approved by the institutional Human Research Ethics Committee (RES-22-00000677L).

### Patient Selection

Consecutive patients referred for elective cardiac electrophysiology (EP) study for evaluation and management of supraventricular tachycardia were considered for inclusion.

Exclusion criteria were: (1) refusal to provide informed consent; (2) pre-existing bundle branch block with QRS duration >120 ms; (3) haemodynamic instability or significant electrolyte disturbances; and (4) the presence of ventricular pre-excitation.

All participants provided written informed consent prior to enrolment.

### Electrophysiology Study Protocol

All procedures were performed in the fasting state (≥4 hours) under light conscious sedation and local anaesthesia. Vascular access was obtained via the right femoral vein, typically with three venous sheaths.

Standard diagnostic catheters were positioned in the coronary sinus (decapolar catheter), His bundle region (quadripolar catheter), and right ventricular (RV) apex (quadripolar catheter). Following completion of the routine EP study and any indicated ablation, pacing protocols were performed at a fixed cycle length slightly faster than the patient’s intrinsic sinus rhythm.

The following pacing conditions were applied:

- RV apical pacing
- His bundle region pacing using low-output pacing (to capture ventricular myocardium) and high-output pacing (to capture both His bundle and ventricular myocardium)
- Coronary sinus pacing (atrial pacing)

These manoeuvres generated different QRS morphologies while maintaining a constant heart rate.

### Electrocardiographic Acquisition

12-lead electrocardiograms (ECGs) were recorded with patients in the supine position using conventional electrode placement. Recordings were obtained at a paper speed of 100 mm/s during each pacing condition at the same cycle length.

Accordingly, four ECG recordings per patient were analysed, each representing a distinct QRS morphology at an identical heart rate.

### ECG Measurements

All ECG measurements were performed digitally using electronic callipers. The R–R interval, QRS duration, and QT interval were measured for each tracing.

QT interval measurements were performed independently by two observers across all leads, and the mean value was used for analysis.

Because all pacing was performed at a fixed cycle length, no heart rate correction of the QT interval was applied.

QRS duration was defined as the interval from the onset of ventricular depolarization to the J point. The QT interval was measured from the onset of the QRS complex (excluding the pacing spike) to the end of the T wave, defined either as the return to the isoelectric baseline or the baseline intersection of a tangent drawn to the steepest downslope of the terminal T wave. U waves were excluded from QT measurements.

### Statistical Analysis

All statistical analyses were performed using IBM SPSS Statistics (version 30; IBM Corp., Armonk, NY, USA). Continuous variables were expressed as mean ± standard deviation, and categorical variables as counts and percentages. Normality was assessed using the Kolmogorov–Smirnov test.

QRS and QT intervals obtained during coronary sinus pacing were considered surrogates for intrinsic conduction. Changes in QT interval (ΔQT) and QRS duration (ΔQRS) were calculated relative to this reference. Pearson correlation analysis was used to assess the relationship between ΔQT and ΔQRS across pacing sites.

To account for repeated measurements within individuals, a linear mixed-effects model was constructed with ΔQT as the dependent variable and ΔQRS as the primary fixed-effect predictor. Pacing site (RV apex, high output pacing and low output pacing from His catheter) was included as a categorical fixed effect, along with an interaction term between ΔQRS and pacing site to assess site-specific differences in slope. A random intercept for each patient was included to account for within-subject correlation. Model assumptions were evaluated using residual diagnostics.

Based on the regression coefficient derived from the mixed-effects model, a linear correction formula was developed: QT corrected = QT paced − N × ΔQRS, where N represents the estimated slope of the relationship between ΔQT and ΔQRS.

Agreement between corrected QT intervals derived from different correction formulas, including the proposed formula and previously published correction methods, and the reference QT interval was assessed using Bland–Altman analysis. For each correction coefficient, the mean difference (bias) and 95% limits of agreement (LoA; mean ± 1.96 × standard deviation) were calculated, together with corresponding confidence intervals. Both absolute and relative differences (difference divided by the mean value) were examined to assess proportional bias. Bland–Altman plots were visually inspected for systematic trends, including heteroscedasticity and proportional error. The performance of each correction coefficient was compared according to: (1) proximity of mean bias to zero; (2) width of the limits of agreement; and (3) absence of proportional bias on visual inspection.

A two-sided p-value <0.05 was considered statistically significant.

## Results

After application of the inclusion and exclusion criteria, a total of 94 patients from Victorian Heart Hospital were enrolled. Baseline characteristics are summarised in Table 1.

**Table 1:** Baseline characteristics.

| Characteristics |  |
| --- | --- |
| Age (years) | 46.74±17.01 (20-80) |
| Gender |  |
| Male sex (N, %) | 43 (45.7%) |
| Female sex (N, %) | 51 (54.3%) |
| BMI, Kg/m<sup>2</sup> | 28.10±5.90 (16.80-53.60) |
| Creatinine (μmol/L) | 76.61±15.61 (51-118) |
| Potassium (mmol/L) | 4.18±0.38 (3.5-5.1) |
| DM (N, %) | 7 (7.5%) |
| LVEF(%) | 62.18±4.75 (50-70) |

BMI: Body Mass Index, DM: Diabetes Mellitus, LVEF: Left Ventricular Ejection Fraction.

Each patient contributed four ECG recordings obtained at an identical pacing rate but with different QRS morphologies. During high-output pacing from the His catheter, ECGs were included only when both His bundle and right ventricular (RV) myocardial capture were present. During low-output pacing, only RV high septal myocardial capture was included. QRS and QT intervals obtained during coronary sinus (CS) pacing were used as surrogates for intrinsic ventricular activation (Table 2). The mean pacing cycle length was 615.4 ± 118.2 ms (range 350–1000 ms).

**Table 2:**
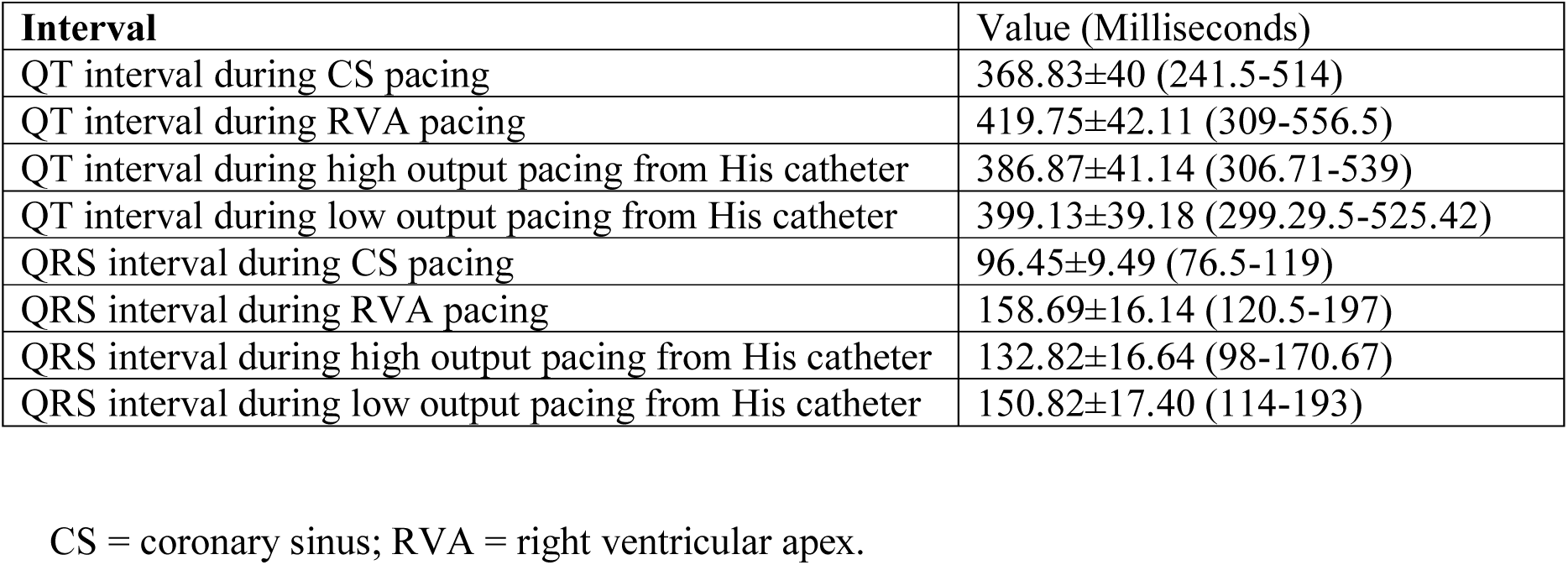
Mean QRS and mean QT intervals during pacing from different locations.

### Site-Specific Analysis

#### Right ventricular apical pacing

RV apical pacing was compared with CS pacing (reference). A total of 94 patients were included. The mean ΔQT was 50.9 ± 17.1 ms, and the mean ΔQRS was 62.1 ± 14.2 ms.

There was a moderate positive correlation between ΔQT and ΔQRS (r = 0.52, p < 0.001). Linear regression analysis demonstrated that ΔQRS was a significant predictor of ΔQT (R^2^ = 0.271, p < 0.001), explaining approximately 27% of the variability in ΔQT. The derived equation was: ΔQT = 12.17 + 0.62 × ΔQRS.

The slope (β = 0.62, SE = 0.107, p < 0.001) indicates that each 10 ms increase in ΔQRS was associated with a 6.2 ms increase in ΔQT. The intercept was not statistically significant (p = 0.077). Residuals were approximately normally distributed, with a standard deviation of 14.6 ms.

### High septal pacing with His capture (high-output pacing)

High-output pacing at the high septum, resulting in combined His and RV capture, was analysed in 93 patients. The mean ΔQT was 18.3 ± 19.1 ms, and the mean ΔQRS was 36.8 ± 17.3 ms.

A strong positive correlation was observed between ΔQT and ΔQRS (r = 0.743, p < 0.001). Regression analysis showed that ΔQRS was a significant predictor of ΔQT (R^2^ = 0.552, p < 0.001), explaining approximately 55% of the variability. The derived equation was: ΔQT = −11.86 + 0.82 × ΔQRS.

The slope (β = 0.82, SE = 0.077, p < 0.001) indicates that each 10 ms increase in ΔQRS resulted in an 8.2 ms increase in ΔQT. The intercept was statistically significant (p < 0.001). Residuals were normally distributed, with a standard deviation of 12.8 ms.

### High septal RV pacing (low-output pacing)

High septal RV pacing without His capture was analysed in 92 patients. The mean ΔQT was 30.3 ± 15.8 ms, and the mean ΔQRS was 54.2 ± 16.1 ms.

A moderate positive correlation was observed (r = 0.524, p < 0.001). Regression analysis demonstrated that ΔQRS significantly predicted ΔQT (R^2^ = 0.275, p < 0.001), explaining approximately 27.5% of the variability. The derived equation was: ΔQT = 2.55 + 0.51 × ΔQRS.

The slope (β = 0.51, SE = 0.088, p < 0.001) indicates that each 10 ms increase in ΔQRS increased ΔQT by approximately 5.1 ms. The intercept was not statistically significant (p = 0.609). Residuals were normally distributed, with a standard deviation of 13.4 ms.

### Overall Relationship Between ΔQRS and ΔQT

Across all pacing sites, ΔQRS and ΔQT demonstrated a consistent positive linear relationship, indicating that increasing QRS prolongation is associated with progressive QT prolongation.

The estimated slopes varied by pacing site (β ≈ 0.51–0.82), suggesting that each 1 ms increase in QRS duration resulted in approximately 0.5–0.8 ms prolongation in QT interval.

### Mixed-Effects Model

To account for repeated measurements within individuals, a linear mixed-effects model was constructed. The model demonstrated strong explanatory power, with a marginal R^2^ of 0.724 and a conditional R^2^ of 0.803, indicating that fixed effects explained 72% of the variability in ΔQT, increasing to 80% when between-subject variability was incorporated. The intraclass correlation coefficient was 0.286.

ΔQRS was a strong independent predictor of ΔQT (β ≈ 0.59, p < 0.001). Pacing site showed a modest but statistically significant overall effect (p = 0.007). However, there was no significant interaction between ΔQRS and pacing site (p = 0.722), indicating that the slope of the ΔQT–ΔQRS relationship remained consistent across pacing locations.

These findings support a unified model in which QT prolongation during ventricular pacing is primarily determined by QRS prolongation, with a stable relationship across different pacing sites.

In a secondary analysis, the high-output His pacing group was excluded, and a second linear mixed-effects model was constructed, with QT interval as the dependent variable, QRS duration as a continuous covariate, pacing location as a fixed factor, and patient as a random effect. QRS duration remained significantly associated with QT interval (F = 35.1, p < 0.001), whereas pacing location itself was not significantly associated with QT interval after adjustment for QRS duration (F = 0.46, p = 0.631). Furthermore, there was no significant interaction between pacing location and QRS duration (F = 0.93, p = 0.394), indicating that the relationship between QRS duration and QT interval was similar across pacing sites.

The absence of a significant interaction in both analyses suggests that the effect of QRS widening on QT prolongation is largely independent of pacing location and supports the application of a single linear correction model across different pacing configurations.

### Bland–Altman Analysis and QT Correction Models

Bland–Altman analysis was performed to assess agreement between corrected QT intervals (QT − N × ΔQRS) and the reference QT interval. A total of 279 paired observations were analysed.

Four candidate correction coefficients (0.55, 0.60, 0.65, and 0.70) were evaluated. Overall, dispersion was similar across models (SD ≈ 16 ms), but systematic bias varied depending on the coefficient (Table 3).

**Table 3:** Agreement between corrected QT intervals and the reference QT interval using four correction models.

| MODEL | Mean Bias (ms) | SD (ms) | Limits of Agreement (ms) | Regression Slope($\beta$) | P value |
| --- | --- | --- | --- | --- | --- |
| 0.55$\times\Delta$QRS | +6.29 | 16.44 | -25.9 to +38.5 | 0.012 | 0.634 |
| 0.60$\times\Delta$QRS | +3.74 | 16.16 | -27.9 to +35.4 | 0.010 | 0.634 |
| 0.65$\times\Delta$QRS | +1.19 | 15.93 | -30.0 to +32.4 | 0.009 | 0.725 |
| 0.70$\times\Delta$QRS | -1.36 | 15.76 | -32.2 to +29.5 | 0.008 | 0.748 |

Regression analysis of Bland–Altman plots demonstrated no evidence of proportional bias for any model.

Increasing the correction coefficient progressively reduced systematic bias, with near elimination at 0.70 × ΔQRS. However, this introduced a slight tendency toward overcorrection. The 0.65 model provided the most balanced performance, achieving low bias with stable variability.

### Effect of Reference QRS Assumption

Because intrinsic QRS duration is not always known in clinical practice, a reference QRS value of 100 ms was used (observed mean 96.45 ± 9.49 ms).

Application of this reference yielded similar dispersion across models, with systematic bias decreasing as the correction coefficient increased. The findings remained consistent with the primary analysis, supporting the use of a correction coefficient in the range of 0.65–0.70, with 0.65 offering the best balance between bias and stability (Supplementary Figure 1).

### Bland–Altman Analysis of previously published QT Correction Formulae

For comparison, QT intervals corrected using two commonly applied formulae, the Bogossian and Talbot correction methods, were compared with the reference QT interval using Bland–Altman analysis. The two correction methods demonstrated distinct agreement characteristics.

The Bogossian formula systematically underestimated the reference QT interval, with a mean bias of −37.1 ms, although it demonstrated relatively narrow limits of agreement. In contrast, the Talbot formula showed substantially lower mean bias (−5.7 ms), indicating closer overall agreement with the reference QT interval.

However, regression analysis demonstrated a small but statistically significant proportional bias for the Talbot correction (β = 0.071, p = 0.033, R^2^ = 0.016), suggesting increasing divergence from the reference QT interval at higher QT values. Conversely, the Bogossian correction showed no significant proportional bias (β = −0.021, p = 0.433, R^2^ = 0.002), indicating more stable agreement across the measured QT range.

Visual inspection of the Bland–Altman plots supported these findings and confirmed the presence of proportional error with the Talbot correction method, whereas the Bogossian formula demonstrated more uniform agreement despite its systematic underestimation of QT duration (Supplementary Figure 2).

### Bland–Altman Analysis After Heart Rate Correction

Bland–Altman analysis demonstrated good agreement between QT intervals corrected using our formula and the reference QT interval after heart rate correction with Bazett’s formula. The mean bias was approximately +5 ms, with 95% limits of agreement ranging from approximately −38 to +48 ms.

Visual inspection of the Bland–Altman plot showed a random distribution of differences around the mean, without increasing dispersion at longer QT intervals. Regression analysis of the differences against the mean QT values revealed no significant proportional bias (β = 0.07, R^2^ = 0.011, p = 0.074), indicating that the performance of the proposed correction remained consistent across the range of QT values (Supplementary Figure 3).

## Discussion

The present study demonstrates that QT prolongation during ventricular pacing is predominantly determined by QRS prolongation and that correction using a fixed proportion of QRS widening substantially improves the accuracy of QT assessment. The principal findings are as follows: (1) a correction coefficient of approximately 0.65-0.70 × ΔQRS minimised systematic bias compared with lower coefficients; (2) improved accuracy was achieved without loss of precision, as variability remained similar across models; and (3) no proportional bias was observed, indicating stable performance across a broad range of QT values.

The mixed-effects analysis represents one of the major strengths of the present study. After accounting for repeated measurements within individuals, ΔQRS remained a strong independent predictor of ΔQT, whereas the interaction between pacing site and ΔQRS was not significant. These findings indicate that although absolute QT values may differ between pacing configurations, the relationship between QRS prolongation and QT prolongation remains remarkably consistent. Accordingly, QRS widening appears to be the principal determinant of apparent QT prolongation during ventricular pacing, supporting the use of a unified correction model across different pacing sites.

Linear regression analyses performed separately for each pacing configuration demonstrated significant associations between ΔQRS and ΔQT at all sites. High-output His bundle pacing showed the strongest correlation, suggesting that during more physiological ventricular activation, QT prolongation is closely related to depolarisation delay itself. In contrast, the weaker association observed during high septal pacing suggests that factors beyond QRS duration may contribute modestly to repolarisation changes in this setting. Nevertheless, the absence of a significant interaction in the mixed model indicates that these differences are insufficient to justify site-specific correction equations.

Based on these findings, we developed and validated a simple QRS-based correction model. A coefficient of 0.65-0.70 provided the best agreement with the reference QT interval, whereas lower coefficients progressively underestimated the contribution of QRS prolongation and resulted in systematic overestimation of QT duration. Conversely, coefficients greater than 0.70 tended to overcorrect and underestimate the QT interval.

Accordingly, the proposed equation:

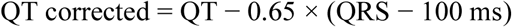

provides a simple and clinically applicable method for estimating the underlying QT interval during ventricular pacing. Importantly, acceptable agreement was maintained using an assumed reference QRS duration of 100 ms, overcoming one of the major limitations of previous correction methods that require knowledge of the patient’s intrinsic QRS duration. This feature makes the proposed approach practical for pacing-dependent patients in whom baseline conduction data are unavailable.

Another important finding was the absence of proportional bias. Bland–Altman analysis demonstrated that the proposed correction performed consistently across the entire QT spectrum, without systematic overcorrection at shorter QT intervals or undercorrection at longer QT intervals. This characteristic is particularly important in patients at increased risk of ventricular arrhythmias, in whom accurate QT assessment is most clinically relevant.

The limits of agreement were similar across all tested models, suggesting that the remaining variability primarily reflects intrinsic biological variation and measurement error rather than deficiencies of the correction method itself. Therefore, optimisation of QT correction appears to depend mainly on reducing systematic bias rather than improving precision.

Previous approaches to QT assessment in patients with broad QRS complexes have relied largely on empirical adjustments or fixed offsets and have seldom been validated using formal agreement analysis. In contrast, the present study derived a correction model directly from the relationship between QRS prolongation and QT prolongation and evaluated its performance using Bland–Altman methodology, which represents the current standard for assessing agreement between measurement techniques.

Comparison with existing correction methods highlighted important limitations of currently available approaches. The Talbot formula demonstrated relatively good overall agreement with the reference QT interval; however, proportional bias was present, indicating that its accuracy varied across the QT spectrum. In contrast, the Bogossian formula demonstrated stable performance across different QT values and showed no evidence of proportional bias, but consistently underestimated the reference QT interval, suggesting systematic overcorrection. Overall, these findings indicate that currently available correction methods may either introduce systematic error or perform inconsistently across different QT ranges.

Taken together, the results of the present study suggest that apparent QT prolongation during ventricular pacing is primarily a consequence of QRS widening and can be corrected using a simple linear adjustment. The proposed formula provides accurate, reproducible, and clinically practical QT estimation across different pacing configurations and may facilitate more reliable assessment of ventricular repolarisation in patients with paced rhythms.

### Clinical Implications

QT correction using a simple linear adjustment based on QRS duration provides a practical and physiologically sound approach for assessing ventricular repolarisation in patients with ventricular pacing or other forms of QRS prolongation. Importantly, the present study demonstrates that an assumed reference QRS duration of approximately 100 ms can be used when intrinsic conduction is unavailable, making the method applicable to pacing-dependent patients and routine clinical practice.

Representative examples from two patients further illustrate the improved agreement between the proposed correction method and the reference QT interval across different pacing configurations (Figures 1 and 2), supporting the robustness of the model at the individual level.

**Figure 1.**
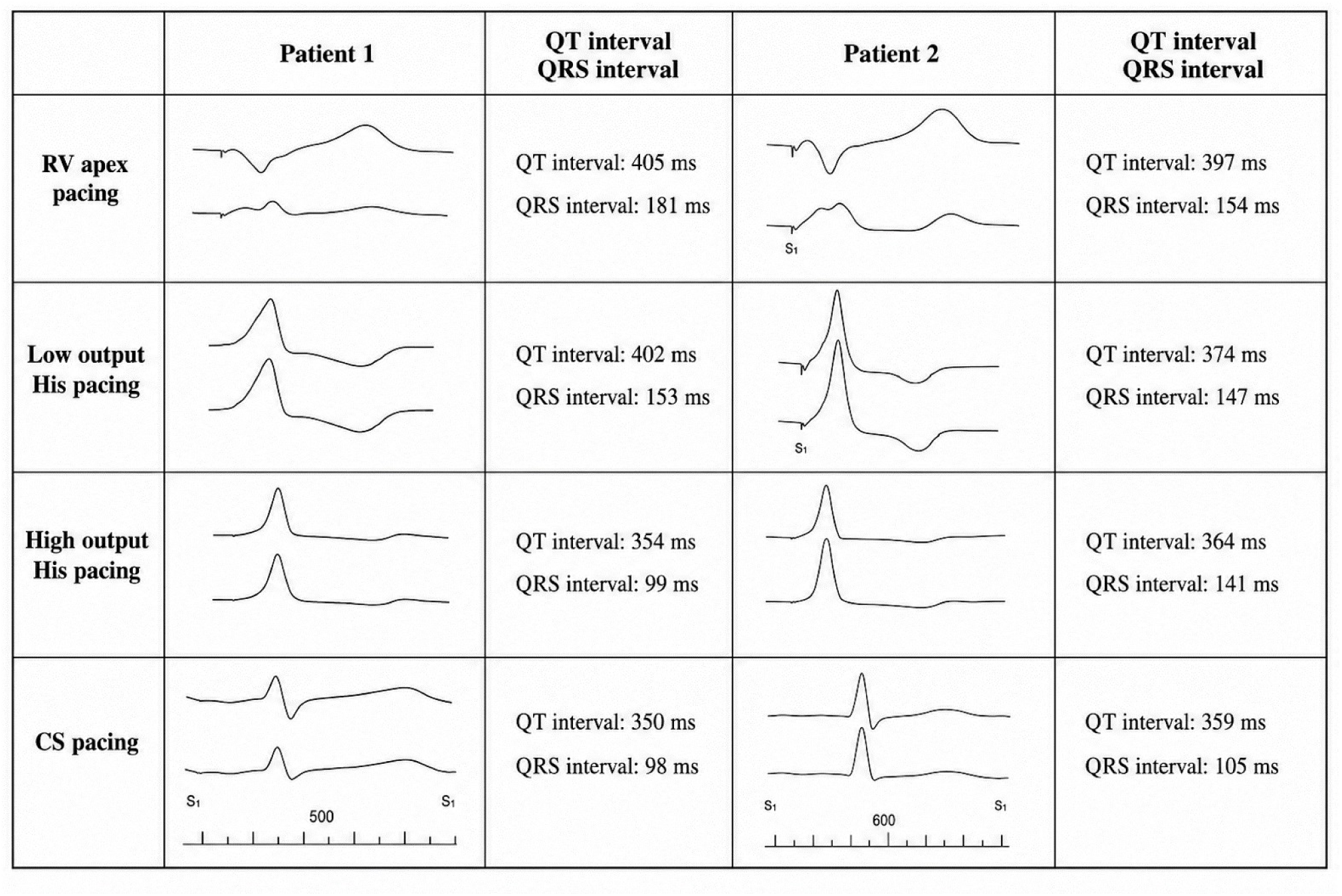
Representative ECGs from two patients demonstrating QT and QRS intervals during right ventricular (RV) apex pacing, low-output His pacing, high-output His pacing, and coronary sinus (CS) pacing. Corresponding QT and QRS interval measurements are shown for each pacing mode in both patients.

**Figure 2.**
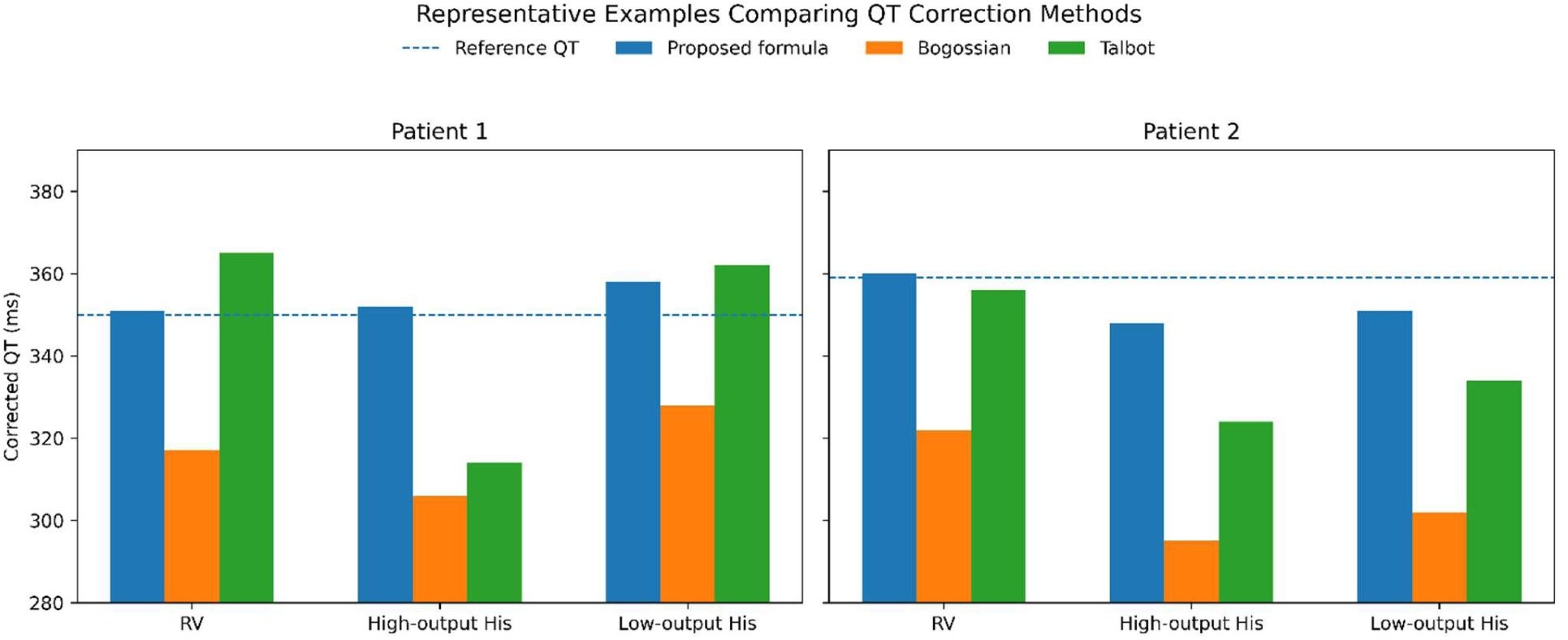
Corrected QT intervals derived using the proposed formula, the Bogossian formula, and the Talbot formula are shown for two representative patients during right ventricular pacing, high-output His pacing, and low-output His pacing. The dashed line represents the reference QT interval.

Nevertheless, the residual variability reflected by the limits of agreement indicates that corrected QT values should be interpreted cautiously in individual patients, particularly when management decisions depend on specific QT thresholds. Therefore, the proposed correction should complement, rather than replace, overall clinical judgement and consideration of other arrhythmic risk factors.

### Limitations

Several limitations of this study should be acknowledged.

First, ventricular pacing was used to reproduce a left bundle branch block (LBBB)-like pattern, which is not fully equivalent to native LBBB physiology. To mitigate this, we included high-output pacing from the His bundle region to achieve combined His and right ventricular capture, providing a more physiological activation pattern.

Second, the persistence of relatively wide limits of agreement suggests that factors beyond QRS duration contribute to QT variability during pacing, including intrinsic repolarisation heterogeneity and measurement variability.

Third, this was a single-centre study, and the findings require external validation in independent cohorts before broader generalisation.

Fourth, the use of a fixed linear correction model may not fully capture more complex electrophysiological relationships across all pacing conditions or patient subgroups.

Fifth, all QT interval measurements were obtained at the same heart rate, and the proposed correction method was not validated across different heart rates or after application of conventional heart rate correction formulas.

Finally, although the proposed correction improves agreement with reference QT values, its direct clinical impact—particularly in risk stratification and clinical decision-making—remains to be established. Prospective studies evaluating the association between the corrected QT interval derived from this model and clinical outcomes are needed to establish its prognostic significance.

### Conclusion

A simple correction using QT corrected = QT − 0.65 × (QRS − 100 ms) provides the best balance between accuracy and precision, with minimal bias and no proportional error. This pragmatic approach offers a clinically applicable method for QT assessment in patients with conduction abnormalities or ventricular pacing.

## Data Availability

NA

## AUTHOR STATEMENT

Mohammad Alasti- Conceptualization, Methodology, Supervision, Data Curation, Writing - Original Draft.

Amin Esmailian- Data collection, Data Curation, Writing - Review & Editing.

Colin Machado- Writing - Review & Editing.

Jagat Adhikari- Data Curation.

Sing Huey Cheng- Data Curation.

Francis Ha- Writing - Review & Editing.

Jeffrey Alison- Writing - Review & Editing.

Andrew Krahn- Writing - Review & Editing.

Hui Chen Han- Writing - Review & Editing.

## Disclosure

The authors declare that there are no financial conflicts of interest or funding sources related to this study.

During the preparation of this work, the authors used artificial intelligence (AI) tools primarily to reassess the statistical analysis results and to assist with editing and manuscript refinement. Following the use of these tools, the authors carefully reviewed and edited the content and accept full responsibility for the accuracy and integrity of the published article

## Aknowledgement

We would like to thank the cardiac physiologists at the Victorian Heart Hospital — Lucy Rudolph, Juliet Young, Sarah Paitry, Thi Bui, Amanda Way, Karlie Chambers and Angela Coumountouros — for their invaluable assistance and support throughout this study.

---

Figure S1: Bland–Altman plots comparing QT intervals corrected using QT-0.65 (QRS-100) and QT-0.70 (QRS-100) formulae with the reference QT interval. The central solid line represents mean bias, while the upper and lower dashed lines indicate the 95% limits of agreement.

Figure S2: Bland–Altman plots comparing QT intervals corrected using Bogossian and Talbot formulae with the reference QT interval. The central solid line represents mean bias, while the upper and lower dashed lines indicate the 95% limits of agreement.

Figure S3: Bland–Altman plots comparing QT intervals corrected using the proposed formula with the reference QT interval after heart rate correction using Bazett’s formula. The solid central line represents the mean bias, while the upper and lower dashed lines denote the 95% limits of agreement.

## References

1. Schwartz PJ, Ackerman MJ. The long QT syndrome: a transatlantic clinical approach to diagnosis and therapy. European heart journal 2013: 34(40): 3109–3116.

2. Roberts JD, Gula LJ. QT-Interval Assessment in Left Bundle Branch Block: Deciphering Normal Within Abnormal. Canadian Journal of Cardiology 2019;35: 802–804

3. Tang JKK, Bennett MT, Rabkin SW. Assessment of QT interval in ventricular paced rhythm: Derivation of a novel formula. Journal of Electrocardiology 2019; 57: 55–62.

4. Yankelson L, Hochstadt A, Sadeh B, Pick B, Finkelstein A, Rosso R, Viskin S. New formula for defining “normal” and “prolonged” QT in patients with bundle branch block. Journal of Electrocardiology 2018;51:481–486.

5. Roberts JD, Gula LJ. QT-Interval Assessment in Left Bundle Branch Block: Deciphering Normal Within Abnormal. Canadian Journal of Cardiology 2019;35: 802–804

6. Erkapic D, Frommeyer G, Crijns HJG, Seyfarth M, Bogossian H. QTc interval evaluation in patients with right bundle branch block or bifascicular blocks. Clin Cardiol. 2020; 43: 957–962.

7. Weipert KF, Bogossian H, Conzen P, Frommeyer G, Gemein C, Helmig I, Chasan R, Eckardt L, Seyfarth M, Lemke B, Zarse M, Hamm CW, Schmitt J, Erkapic D. Application of the Bogossian formula for evaluation of the QT interval in pacemaker patients with stimulated left bundle branch block. Clinical Research in Cardiology (2018) 107:1033–1039.

8. Fredholm K, Højlund M, Bjerregaard MR, Nielsen JC, Fink-Jensen A, Correll CU. How to Estimate QT Interval in Patients with Left or Right Bundle Branch Block Journal of Clinical Psychopharmacology 2021; 41 (3): 323–326.

9. Tabatabaei P, Keikhavani A, Haghjoo M, Fazelifar A, Emkanjoo Z, Zeighami M, Bakhshandeh H, Ghadrdoost G, Alizadeh A. Assessment of QT and JT Intervals in Patients With Left Bundle Branch Block. Res Cardiovasc Med. 2016 May; 5(2): e31528.

10. Bogossian H, Frommeyer G, Ninios I, Pechlivanidou E, Hasan F, Nguyen QS, Mijic D, Kloppe A, Karosiene Z, Margkarian A, Bandorski D, Schultes D, Erkapic D, Seyfarth M, Lemke B, Eckardt L, Zarse M. A new experimentally validated formula to calculate the QT interval in the presence of left bundle branch block holds true in the clinical setting. Ann Noninvasive Electrocardiol. 2017;22:e12393.

11. Wang B, Zhang L, Cong P, Chu H, Liu Y, Liu J, Surkis W, Xia Y. A New Formula for Estimating the True QT Interval in Left Bundle Branch Block. Cardiovasc Electrophysiol 2017; 28: 684–689.

12. Abu-El-Haija B, J Paul Mounsey AP. QT interval correction in LBBB: Improving the accuracy of an imperfect measurement J Cardiovasc Electrophysiol.2017;28:690–691.

13. Frommeyer G, Bogossian H, Pechlivanidou E, Conzen P, Gemein C, Weipert K, Helmig I, Chasan R, Johnson V, Eckardt L, Hamm CW, Seyfarth M, Lemke B, Zarse M, Schmitt J, Erkapic D. Applicability of a Novel Formula (Bogossian formula) for Evaluation of the QT-Interval in Heart Failure and Left Bundle Branch Block Due to Right Ventricular Pacing. PACE 2017; 40:409–416.

14. Talbot S. QT interval in right and left bundle-branch block. British Heart Journal 1973; 35, 288–291.

15. Rautaharju PM, Zhang ZM, Prineas R, Heiss G. Assessment of Prolonged QT and JT Intervals in Ventricular Conduction Defects. Am J Cardiol 2004;93:1017–1021

16. Funk MC, Cates KW, Rajagopalan A, Lane CE, Lou J. Assessment of QTc and Risk of Torsades de Pointes in Ventricular Conduction Delay and Pacing: A Review of the Literature and Call to Action. Journal of the Academy of Consultation-Liaison Psychiatry 2021:62:501–510.

17. Sriwattanakomen R, Mukamal KJ, Shvilkin A. A novel algorithm to predict the QT interval during intrinsic atrioventricular conduction from an electrocardiogram obtained during ventricular pacing. Heart Rhythm 2016;13:2076–2082.

18. Viskin S, Uri Rosovski U, Sands AJ, MPhil, Chen E, Kistler PM, Kalman JM, Chavez LR, Torres PI, Cruz F FES, Centurión OA, Fujiki A, Maury P, Chen X, Krahn AD, Roithinger F, Zhang L, Vincent M, Zeltser D. Inaccurate electrocardiographic interpretation of long QT: The majority of physicians cannot recognize a long QT when they see one. Heart Rhythm 2005; 2:569–574.

